# Three-Month Interim Analyses of Repeated Low-Level Red-Light Therapy in Myopia Control in Schoolchildren: A Multi-Ethnic Randomized Controlled Trial

**DOI:** 10.1101/2024.03.16.24304399

**Authors:** Nellie Deen, Zhuoting Zhu, Ziyi Qi, Yuri Yin-Moe Aung, Gabriella Bulloch, Di Miao, Mingguang He

## Abstract

**Purpose:** To assess the efficacy and safety of repeated low-level red-light (RLRL) therapy in controlling myopia progression among multi-ethnic school-aged children. This report focuses on 3-month interim analysis.

**Design:** Multi-ethnic, parallel controlled randomized trial

**Participants:** A total of 34 children aged 8-13 years with myopia of cycloplegic spherical equivalent (SE) of −0.50 to – 5.00 (inclusive) diopters (D), astigmatism of 2.50 D or less, anisometropia of 1.50 D or less, and monocular best-corrected visual acuity (BCVA) of 20/20 or better were enrolled.

**Methods:** Participants were randomly assigned to the RLRL group (n = 16) or the single-vision spectacles (SVS) group (n = 18). RLRL therapy was administered twice daily on weekdays for 3-minute sessions, while the SVS group continued routine activities. Assessments were scheduled at baseline and follow-up visits at 1, 3, 6, and 12 months, with compliance monitoring and safety assessments throughout.

**Main Outcome Measures:** The primary outcome and a key secondary outcome included axial length (AL) change and cycloplegic spherical equivalent (SE) change.

**Results:** A total of 31 (91.2%) participated in the 3-month follow-up visit. The RLRL group demonstrated a significant shortening in AL (−0.07 ± 0.07 mm) compared to the SVS group (0.03 ± 0.05 mm, P<0.001). Similarly, SE progression was hyperopic shift in the RLRL group (0.26 ± 0.14 D) while the SVS group exhibited a myopic shift (−0.03 ± 0.38 D, P=0.009). No severe adverse events were reported.

**Conclusions:** The 3-month interim analysis shows that the efficacy of RLRL therapy in controlling myopia progression among multi-ethnic children is comparable to, or even better than, that idenfied in Chinese patients in previous trials.

## Introduction

Myopia has emerged as a public health concern with a rising prevalence worldwide, particularly among school-aged children^1-3^. Further progression towards severe myopia, termed as high myopia, is associated with an elevated risk of irreversible blindness-inducing conditions such as myopic maculopathy, glaucoma, and retinal detachment^4,5^. Consequently, effectively controlling myopia progression is critically important to safeguard childhood vision, preserving eye health and future quality of life.

Among the diverse interventions that have been explored for myopia control, such as increased time outdoors^6-8^, orthokeratology and low-dose atropine^9, 10^, low-level red-light therapy (RLRL)^11-16^ has garnered attention for its stronger efficacy in controlling the myopia progression. In contrast to strategies relying on increased outdoor exposure thereby increasing ambient light exposure, RLRL utilizes a device emitting 650-nm visible red light onto the fundus, providing higher light energies for shorter durations. A previous pivotal 12-month multicenter trial conducted in China demonstrated the efficacy and safety of RLRL therapy in controlling myopia progression, reporting a significant reduction in axial length (AL) elongation and spherical equivalent (SE) progression compared to single-vision spectacles (SVS), with no documented functional or structural damage^13^. Another 12-month trial in China then explored the preventative potential of RLRL therapy in premyopic children, revealing a 33.4% reduction in myopia incidence relative to the control group^14^.

While promising outcomes have also been observed in several other Chinese studies, the applicability of RLRL across diverse ethnicities remains unknown. Therefore, in order to establish the generalizability of this intervention as a novel and globally applicable solution for controlling myopia in school-aged children, a single-blind, multi-ethnic, parallel-group randomized controlled trial (RCT) was designed to validate the efficacy and safety of RLRL therapy in a multi-ethnic population. This manuscript presents the interim 3-month findings of this study, offering a preliminary understanding into the effects of RLRL therapy on AL and SE in an ethnically diverse cohort of schoolchildren.

## Methods

### Study Design

This prospective, multi-ethnic, parallel controlled randomized trial was conducted in Melbourne, Australia. Children were enrolled through two pathways: retrospective electronic medical record data within the past 12 months or on-site recruitment from refraction clinics and optometry centers between August 2022 and November 2023. All examinations at baseline and follow-up visits were conducted by assigned study optometrists, following the trial protocol. For overall trial duration, examinations were conducted at baseline and at the 1-, 3-, 6-, and 12-month follow-up visits, with the trial scheduled for completion in December 2024.

The study protocol was approved by the Ethics Committee of The Royal Children’s Hospital (Identifier, HREC 82296), and adhered to the tenets of the Declaration of Helsinki. Written informed consent was obtained from legal guardians of the enrolled children. An independent data and safety monitoring committee (DSMC) ensured unbiased oversight of the clinical trial, with all participants were covered by a three-year research insurance indemnity scheme. The study was registered on the Australian New Zealand Clinical Trials Registry (ANZCTR) website (Identifier, ACTRN12622000526774p).

### Eligibility Criteria

Eligible participants were children aged 8-13 years with myopia, defined as a cycloplegic SE of −0.50 to –5.00 (inclusive) diopters (D), astigmatism of 2.50 D or less, anisometropia of 1.50 D or less, and monocular best-corrected visual acuity (BCVA) of 20/20 or better. These criteria have been used in previous trials^13^. Chinese participants comprised less than 10% of the overall recruitment. Participants had to be willing to participate in all required activities of the study and accept random allocation of grouping. Children were excluded if they had strabismus, binocular vision abnormalities, other ocular abnormalities, any systemic diseases, or a previous history of any myopia interventions other than single vision lenses. Participants were also excluded if the optometrist thought they had any contraindications that made them unsuitable for enrolment. Participants were to withdraw and stop the treatment in the occurrence of any severe adverse event (a sudden loss of vision > two lines, or a scotoma developed in the centre of the visual field) and annual refraction progression > 1.50 D.

### Randomization and Masking

Randomization schedules using a pseudorandom number generator were stratified by ethnic background. Within each ethnic background, eligible participants were randomized in a 1:1 ratio to receive either RLRL therapy in conjunction with SVS in the intervention arm or SVS correction only in the control arm. The randomization process was based on the sequentially numbered, opaque, sealed envelope. The randomization list in the system was pre-generated by a statistician who had no prior contact with any study investigators. The study identification, name of participant and group allocation assigned were frozen in the system where further changes were not allowed.

Group allocation was concealed from study researchers until the baseline visit was completed for each enrolled participant. Given the nature of the intervention, participants were aware of their study allocation. However, technicians, study optometrists, and statisticians, were all masked.

### Intervention

The study employed a semi-conductor laser product (Eyerising International Pty Ltd, Melbourne, Australia) emitting low-level red light at 650 ± 10 nm, with a power output of 2.00 ± 0.50 mW and a laser spot diameter at the observation port of 10 mm ± 2 mm. The device was tested and certified as group 1 in the ANSI Z80.36-2021 meaning no potential light hazard^17^. The device, equipped with internet connectivity and automated diary functions, administered treatment sessions following a specific schedule: 3-minute sessions twice a day, with a minimum 4-hour interval, 5 days a week. Parents received thorough instructions and supervised each session.

Children in the intervention group, alongside their routine activities and wearing single vision spectacles, underwent the therapy regimen under parental supervision. They were reminded to keep their eyes open during the 3-minute sessions and were discouraged from leaving the device. Conversely, the control group received no red-light therapy, continuing routine activities with single vision spectacles for myopia correction. No other myopia interventions were administered to either group for the duration of their participation in the trial.

### Intervention Compliance Monitoring

Participants were required to log in with unique accounts and passwords before initiating each treatment session. The device was connected to a central system via the internet, logging device use. To ensure adherence, project staff conducted weekly reviews of treatment compliance and usage statistics. Parents or guardians of participants with <80% compliance received text reminders to promote compliance.

### Data Collection

Axial length (AL) was measured before cycloplegia using the IOLMaster (Carl Zeiss 500, Meditec, Oberkochen, Germany) and averaged until the maximum error did not exceed 0.10 mm.

Cycloplegia was induced using 1 drop of 0.5% Alcaine (Alcon, Puurs, Belgium) followed by 3 drops of 1% cyclopentolate (Alcon, Puurs, Belgium) to each eye at 0, 5, and 20 minutes. Full cycloplegia was defined as a pupil diameter of at least 6 mm and an absent pupillary reaction to light. Refraction data were measured using an autorefractor (KR-8800, Topcon, Tokyo, Japan) 3 times after cycloplegia and averaged until the desired precision (spherical and cylindrical power < 0.25 D, axis < 5 degree) was achieved. The spherical equivalent was calculated by the sum of spherical power and half of cylindrical power.

Other ocular biometric parameters including anterior chamber depth (ACD), corneal curvature (CC), and white-to-white (WTW) corneal diameter were measured at the same session as AL measurement on each eye before cycloplegia by IOLMaster and were averaged once their desired precisions were achieved.

Uncorrected visual acuity (UCVA) and best-corrected visual acuity (BCVA) were assessed from 4 meters by trained optometrists using the Early Treatment Diabetic Retinopathy Study (ETDRS) visual acuity chart (Precision Vision, Villa Park, Illinois, USA).

Optical coherence tomography (OCT) scans were obtained from swept-source OCT (SS-OCT, DRI-OCT Triton, Topcon, Tokyo, Japan) under radiographic scanning (radial 12 mm × 9 mm) centered at the fovea. The quality of the scans was indicated by an automated display mode.

### Outcomes

Study outcomes focus on efficacy and safety of RLRL therapy in achieving myopia control. The primary outcome was the AL change at the follow-up compared to baseline in both groups. Secondary outcomes included changes in cycloplegic SE (defined as myopia progression), other ocular biometric parameters including ACD, CC, and WTW corneal diameter, visual acuity (VA) at the follow-up visit, compared to baseline in both groups. The trial also considered self-reported adverse events (AEs) and OCT scans.

### Adverse Events

Participants who underwent at least 1 session of treatment were analyzed for safety. A questionnaire on adverse events (AEs) post-intervention was administered at each follow-up and any unexpected visits for intervention group participants. Participants and their parents/guardians were queried about AEs including, but not limited to, short-term glare, flash blindness, and afterimages. Additionally, two ophthalmologists independently reviewed all OCT scans to identify any possible structural damages.

### Sample Size

Using the outcomes from the previous study, where the mean difference in axial elongation between the RLRL treatment and SVS control groups was 0.26 mm^13^, and assuming a standard deviation of 0.3 mm for these changes (based on typical clinical trial variability for such measurements^13, 14^), we aim to detect a similar difference with a statistical power of 80% and a significance level of 5%. The calculations suggest that a total of 42 participants is necessary to achieve these objectives, with 21 participants in each group (RLRL treatment group and SVS control group).

### Statistical Analysis

The interim analysis included data from all participants attending the 3-month follow-up visit, with the right eye serving as the representative outcome. Group comparisons utilized the χ2 test for categorical outcome such as sex and the unpaired t-test for continuous outcomes with a normal distribution such as AL and SE; nonparametric methods were used if otherwise. Statistical analyses were conducted using Stata (StataCorp, Stata Statistical Software: Release 14; College Station, TX, USA). All statistical tests were two-sided and performed at the 5% significance level unless otherwise noted.

## Results

### General information

Between August 2022 and November 2023, children with myopia (n = 41) were recruited and assessed for eligibility in Melbourne. 34 children (82.9%) met the inclusion criteria, with 16 randomly assigned to the RLRL group and 18 to the SVS group. Among them, the ethnic distribution was as follows: Indian (6); Caucasian (5); Somalian (5); Pakistan (4); Lebanese (3); Chinese (1), Vietnamese (1), Bangladeshi (1), Burmese(1), Indigenous (1), Iraq (1), Ethiopia (1), Filipino (1), Japanese (1) others (2). Figure 1 summarizes participant flow from enrolment to the 3-month follow-up visit. Among the 34 children, 31 (91.2%) participated in the 3-month study, comprising 16 children (100%) in the RLRL group and 15 children (83.3%) in the SVS group.

**Fig 1.**
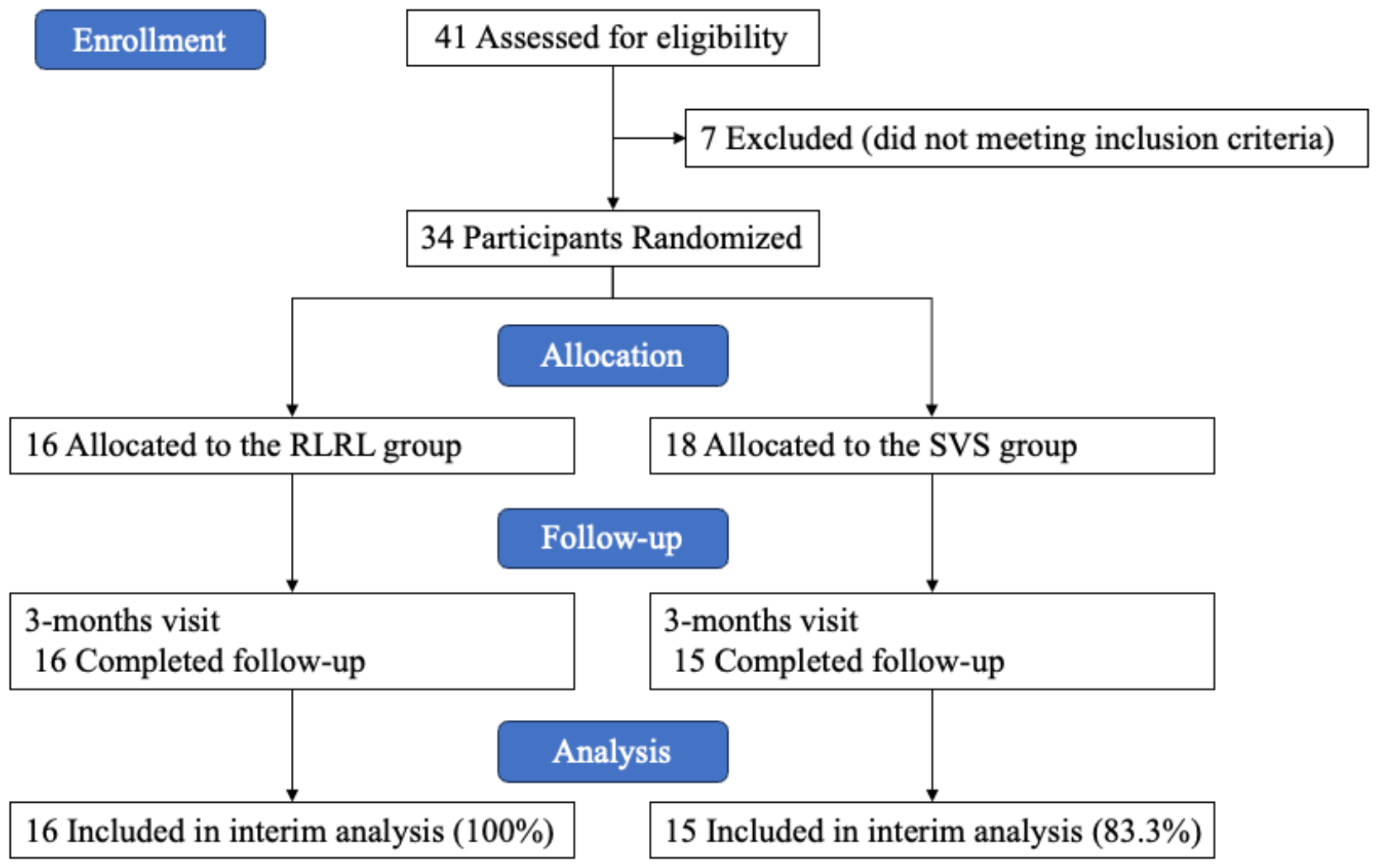
Flowchart of the Study. RLRL, repeated low-level red-light; SVS, single-vision spectacle.

### Baseline Characteristics

The mean age and proportion (%) of male children were comparable between the RLRL and SVS groups (11.52 ±1.55 years vs. 11.94 ± 1.39 years; male sex, 56.25% [n =9] vs. 55.56% [n =10]). There were no significant differences observed between the groups in terms of baseline AL and SE, as presented in Table 1.

**Table 1.** Demographics and Baseline Characteristics of Study Participants.

| Characteristics | RLRL (N=16) | SVS (N=18) | *P* value |
| --- | --- | --- | --- |
| Age (yrs) |  |  |  |
| Mean±SD | 11.5±1.55 | 11.9±1.39 | 0.407* |
| Sex, No. (%) |  |  |  |
| Male | 9 (56.25) | 10 (55.56) | 0.968<sup>+</sup> |
| Female | 7 (43.75) | 8 (44.44) |  |
| AL (mm) |  |  |  |
| Mean±SD | 24.2±0.69 | 24.6±0.75 | 0.153* |
| SE (D) |  |  |  |
| Mean±SD | -2.18±0.86 | -2.24±1.06 | 0.867* |

RLRL, repeated low-level red-light; SVS, single-vision spectacle; AL, axial length; SE, spherical equivalent; D, diopter; mm, millimeter.
Data are presented as mean ± standard deviation, number (%).
\*Independent-samples t test. <sup>+</sup>Chi-square test.

### Primary Outcome

The primary outcome of AL change was evaluated at 1 month and 3 months. In the RLRL group, a statistically significant AL shortening was observed at 1 month (−0.05 ± 0.04 mm, P<0.001) and further shortening was observed at 3 months (−0.07 ± 0.07mm, P<0.001; Table 2, Fig 2A). Conversely, the SVS group showed a modest AL elongation at 1 month (0.01 ± 0.04 mm) and 3 months (0.03 ± 0.05 mm; Table 2, Fig 2A).

**Table 2.** Changes in Axial Length and Cycloplegic Spherical Equivalent from Baseline to 1- and 3-Months.

| Visit | RLRL | SVS | *P* value* |
| --- | --- | --- | --- |
| Primary outcome |  |  |  |
| Change of AL, mean±SD, mm | | | |
| 1 month | -0.05±0.04 | 0.01±0.04 | <0.001 |
| 3 months | -0.07±0.07 | 0.03±0.05 | <0.001 |
| Secondary outcome |  |  |  |
| Change of SE, mean±SD, D |  |  |  |
| 1 month | 0.10±0.11 | -0.01±0.23 | 0.127 |
| 3 months | 0.26±0.14 | -0.03±0.38 | 0.009 |

RLRL, repeated low-level red-light; SVS, single-vision spectacle; AL, axial length; SE, spherical equivalent; D, diopter; mm, millimeter.
Data are presented as mean ± standard deviation.
\*Independent-samples t test.

**Fig 2.**
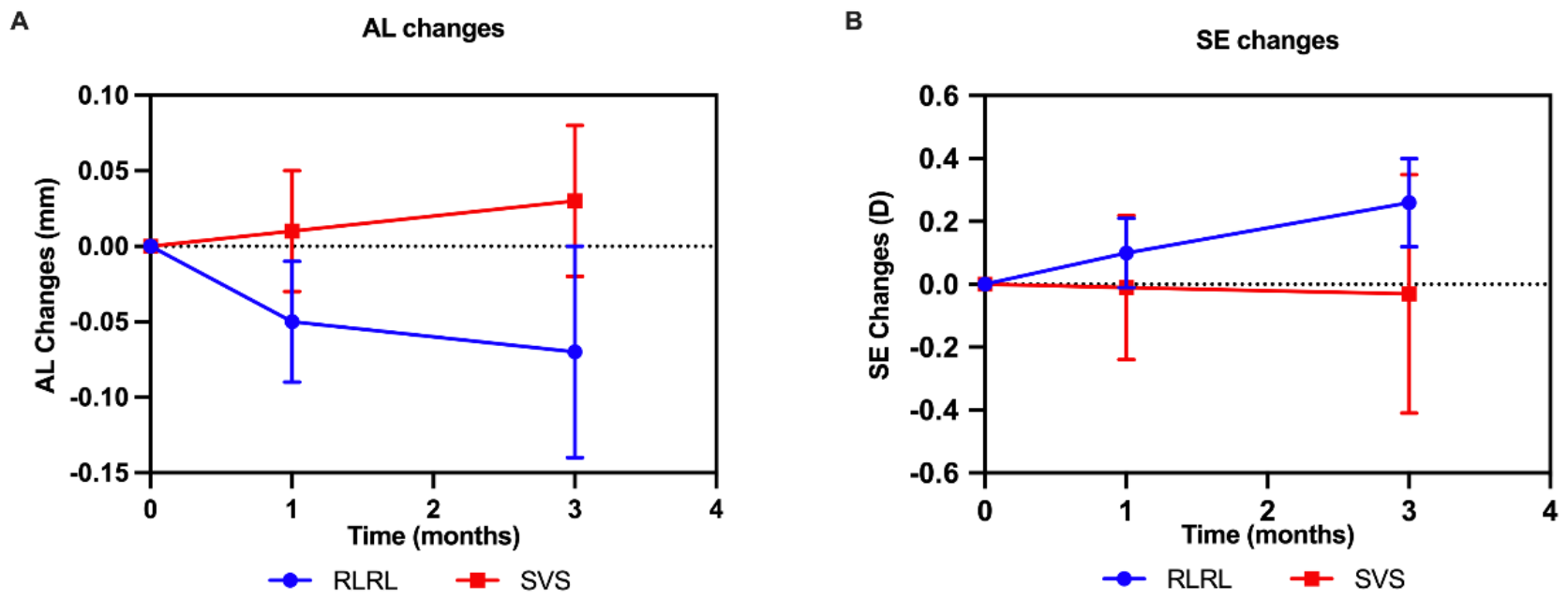
Line Graphs Showing the Changes in Axial Length (A) and Cycloplegic Spherical Equivalent (B) from Baseline to 3-Months at Each Time Point. AL, axial length; SE, spherical equivalent; RLRL, repeated low-level red-light; SVS, single-vision spectacles; D, diopter; mm, millimeter. The error bars represent standard deviation (SD).

### Key Secondary Outcome

At 1 month, there was no significant difference in SE change between the RLRL group (0.10 ± 0.11 D) and the SVS group (−0.01 ± 0.23 D) with a p value of 0.127 (Table 2, Fig 2B). However, at 3 months, the RLRL group demonstrated a reversal in SE (0.26 ± 0.14 D), whereas the SVS group exhibited a significant decrease in SE (−0.03 ± 0.38 D, p = 0.009; Table 2, Fig 2B).

### Adverse Events

As of the 3-month follow-up visit, no severe adverse events (cases of vision loss by two lines or scotoma) were reported. Additionally, there were no indications of glare, flash blindness, or afterimages following the treatment. All children in both the RLRL and SVS groups achieved a BCVA of 20/20. OCT data analysis revealed no structural damage in participants who underwent RLRL therapy. One child in the RLRL group discontinued the treatment due to headache.

## Discussion

In the three-month interim analysis of this 12-month, multi-ethnic, randomized clinical trial, RLRL therapy demonstrated AL shortening of 0.07 mm and a reversal of SE (hyperopic shift) by 0.26 D compared to baseline. In contrast, the SVS group exhibited 0.03 mm AL elongation and 0.03 D SE progression. No severe adverse events were observed. The overall retention rate was satisfactory, with 91.2% of participants completing the 3-month follow-up.

### AL Shortening and Reversal of SE Progression

We previously considered myopia as a progressive and irreversible ocular disease ^18^. The interim analysis of this study showed significant AL shortening at 1 (−0.05mm) and 3 (−0.07mm) months, along with reversal of SE at the 1- (0.10D) and 3-month follow-up (0.26D) in the RLRL group. In contrast, the SVS group exhibited typical AL elongation and SE progression during the same period. This suggests that RLRL therapy may have a mitigating effect on AL elongation.

Studies on RLRL therapy in China have demonstrated sustained choroidal thickening throughout the one-year treatment period^15, 16, 19^. Axial myopia has been shown to be the result of the upregulation of hypoxia-induced factor-1α (HIF-1α) expression due to scleral hypoxia, leading to fibroblast-to-myofibroblast differentiation, scleral remodeling, and decreased mechanical strength^20, 21^. Therefore, we speculate that RLRL therapy may inhibit tthe myopia progression by increasing choroidal blood flow, promoting choroidal thickening and improving oxygen and nutrient supply to the avascular sclera nearby.

### Comparison between Australian and Chinese results

There have been various studies into the use of RLRL within Chinese cohorts. A previous study showed that RLRL can significantly reduce myopia progression (−2.87 ± 1.89 D vs -3.57 ± 1.49 D, p < 0.001) and AL elongation (−0.06 ± 0.19 mm vs 0.26 ± 0.15 mm, p < 0.001) within 9 months^15^. A double-blind RCT with 112 Chinese children aged 7-12 years found that AL elongation (0.01 mm vs. 0.39mm,, p < 0.05) and SE progression (0.05D vs. -0.64D, p < 0.05) in the RLRL group were significantly lower compared to the control group^12^. Beyond slowing myopia progression, RLRL has also been demonstrated to prevent myopia onset^14^. In a 12-month RCT involving 139 children with premyopia, the RLRL group exhibited a 54.1% reduction in myopia incidence compared to the control group.

This Melbourne-based study however provides new evidence into the applicability of RLRL therapy across different ethnicities, extending beyond the positive outcomes observed in Chinese children. Compared to a previous 12-month RCT in Chinese children with similar age and refractive errors ^13^, the Chinese RLRL group showed AL changes at 1- and 3-months of -0.04mm and -0.01mm respectively, with some children experiencing AL elongation at 3 months. In comparison, the Australian RLRL group demonstrated AL elongation at 1- and 3-months of -0.05mm and -0.07mm (Fig 3a), indicating a more prolonged inhibitory effect of RLRL on AL elongation in Australian children of diverse ethnic backgrounds. Similar trends were observed in SE changes. Children in the Australian RLRL group exhibited a more pronounced effect in suppressing SE progression (0.26D vs 0.07D at 3 months, p < 0.05, Fig 3b).

**Fig 3.**
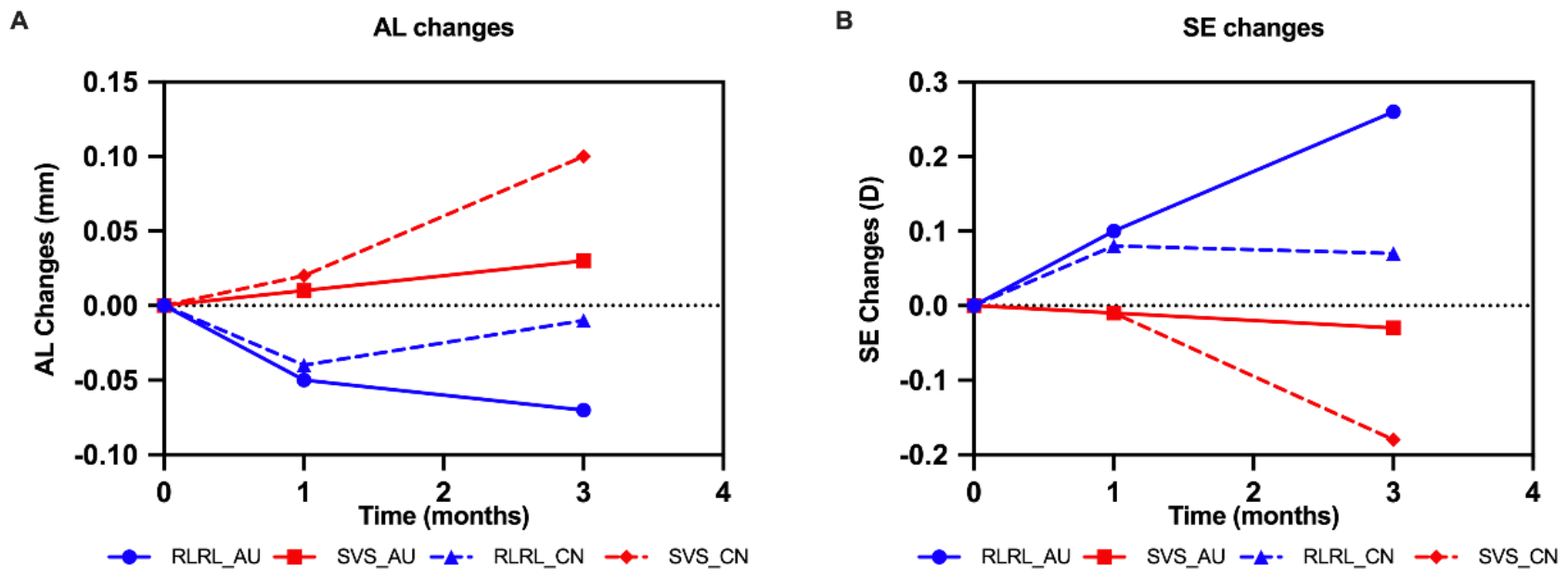
Comparative Analysis of Axial Length (A) and Cycloplegic Spherical Equivalent (B) Changes Between Australian and Chinese Results from Baseline to 3-Months at Each Time Point. The Chinese data were derived from the multicentre study conducted by Jiang et al^13^. AL, axial length; SE, spherical equivalent; RLRL, repeated low-level red-light; SVS, single-vision spectacles; D, diopter; mm, millimeter. The error bars represent standard deviation (SD).

While previous Chinese studies have validated the efficacy of RLRL therapy, our multi-ethnic results confirm that these benefits are not limited to a specific ethnicity. This underscores the potential global applicability of RLRL therapy as a strategy for myopia control.

### Efficacy in Comparison with Other Treatments

Currently, commonly used clinical interventions, such as low-concentration atropine, orthokeratology, defocus-designed spectacle lenses, and soft contact lenses, show variable efficacy in slowing myopia progression, ranging from 30% to 80%^22-24^. Our study revealed that after three months of intervention, RLRL significantly reduced myopic shifts in AL and SE compared to the control group (differences of 0.10mm and 0.29 D, respectively).

In comparison, a Taiwanese study found that increased time outdoors reduced AL elongation by 0.05 mm and SE progression by 0.12 D after one year^7^. In the Low-Concentration Atropine for Myopia Prevention (LAMP2) study in Hong Kong, at four months, the mean SE changes were 0.16D in the 0.05% atropine group, -0.01D in the 0.01% atropine group, and -0.15D in the placebo group. Mean AL elongations were also 0.08mm in the 0.05% atropine group, 0.11mm in the 0.01% atropine group, and 0.16mm in the placebo group^10^. Additionally, a study directly comparing RLRL and orthokeratology lens over six months suggested that RLRL therapy might be slightly more effective, with both significantly reducing AL elongation (RLRL: -0.06±0.15 mm, orthokeratology: 0.06±0.15 mm)^11^. While direct comparisons between these studies are challenging due to differences in measurements and outcomes, RLRL efficacy appears to be at least competitive with other treatment modalities.

### Safety

No serious adverse events were observed after three months of treatment. All participants achieved a BCVA of 20/20. There were no reports of glare, flash blindness, or afterimages, further supporting the safety of RLRL therapy. No other structural changes were identified in this study, consistent with other RLRL studies^12-14^. Existing safety evidence suggests that red light is a low-risk therapy, and it has been used for myopia treatment in China for a few years without reports of long-term adverse effects^16,25, 26^.

### Limitations and future directions

This study has certain limitations. Firstly, the relatively short duration of the interim analysis highlights the necessity for long-term follow-up to evaluate the sustained safety, efficacy and rebound effects of RLRL treatment. Secondly, the observed therapeutic effects of red light in controlling myopia progression are specificto the parameters used in this study. The threshold for the effectiveness of red light, optimal dosage, safety thresholds, and the underlying mechanisms require further exploration. Thirdly, the sample size in this study was relatively small, so a larger sample size would enhance the statistical robustness. Fourthly, the lack of masking raised the possibility of overestimating the effect of RLRL, suggestinf future research should incorporate a sham device as a placebo. Lastly, based on the current analysis, we are unable to describe whether there is regression or rebound in myopia control effects after the cessation of RLRL. Previous studies have reported a slight myopic rebound after cessation of RLRL therapy^12, 16, 25^, and future long-term follow-up is needed to further explore the optimal intervention duration and after-effects.

## Conclusion

This interim analysis substantiates RLRL therapy’s effectiveness and safety in controlling myopia progression. The observed AL shortening, reversal of SE progression, and consistent outcomes across diverse ethnicities suggest the potential of RLRL therapy as a globally applicable solution for myopia control. Anticipating the one-year results, we look forward to further evaluate RLRL as an alternative treatment approach.

## Data Availability

Data will be made available upon reasonable request. To request access, please contact the corresponding author.

## Abbreviations and Acronyms

RLRL=: repeated low-level red-light
SVS=: single-vision spectacles
SE=: spherical equivalent
D=: diopters
AL=: axial length
VA=: visual acuit
BCVA=: best-corrected visual acuity
UCVA=: uncorrected visual acuity
RCT=: randomized Controlled Trial
DSMC=: data and safety monitoring committee
ACD=: anterior chamber depth
CC=: corneal curvature
WTW=: white-to-white
OCT=: optical coherence tomography
AE=: adverse event

## Acknowledgements

The authors thank the participants who were involved in this trial, without whom the trial would not have been possible, and optometrists from Australia College of Optometry, who provided valuable input throughout the study: Marianne Coleman, Diba Rezazadeh, Dzung Tran, Christa Sipos-Ori, and Zeinab Fakih.

